# A Promising Biomarker for Predicting Cardiovascular Risk in Asymptomatic Familial Hypercholesterolemia Patients From The SAFEHEART Study

**DOI:** 10.1101/2025.11.03.25339452

**Authors:** Rafael M. Molero-Payan, Oriol Alberto Rangel-Zuñiga, Alberto Díaz-Cáceres, Ovidio Muñiz-Grijalvo, José Luis Díaz-Díaz, Daniel Zambón, Rodrigo Alonso, Leopoldo Perez de Isla, José López-Miranda, Pablo Pérez-Martínez, Pedro Mata, Francisco Fuentes

**Author notes:** Correspondence should be addressed to Prof. Francisco Fuentes and Oriol Alberto Rangel Zuñiga. Lipids and Atherosclerosis Unit. Department of Medicine. Reina Sofia University Hospital. University of Cordoba. Av. Menendez Pidal s/n. 14004 Cordoba, Spain. Rafael M. Molero-Payan and Oriol Alberto Rangel-Zuñiga contributed equally and are cofirst authors. **Clinical Trial Registration:** ClinicalTrials.gov, number NCT02693548.

## Abstract

**Background:** Familial hypercholesterolemia (FH) is an autosomal dominant disorder associated with a markedly increased risk of premature atherosclerotic cardiovascular disease (ASCVD). Genetic and molecular factors, including microRNAs (miRNAs), may serve as biomarkers to improve disease risk prediction. miRNAs are key regulators of biological processes involved in cardiovascular pathology. This study aimed to identify miRNAs that, combined with clinical variables, enhance ASCVD prediction in asymptomatic FH patients from the SAFEHEART cohort.

**Methods:** A total of 400 subjects diagnosed with FH were included. Coronary calcium score (Agatston score) was measured by computed tomography and stratified into three risk categories (0–99, 100–299, >300). Circulating plasma miRNA levels were quantified using the OpenArray platform. Statistical analyses characterized baseline features and assessed predictive performance using ROC curve analysis. Functional enrichment was explored through the MIENTURNET database.

**Results:** Individuals carrying the null allele for the LDL-R gene showed significantly higher plasma *miR-1* levels than those carrying the defective allele (p=0.042). Likewise, subjects with an Agatston score of 0–99 exhibited higher *miR-1* levels than those with scores >300 (p=0.025). A predictive model based on clinical variables including age, sex, hypertension, smoking, BMI, LDL burden, and lipoprotein(a) achieved an AUC of 0.846 for ASCVD prediction. Incorporating *miR-1* increased the AUC to 0.878, representing a statistically significant improvement (DeLong test, p=0.005).

**Conclusions:** *miR-1* emerged as a promising biomarker for ASCVD risk prediction in FH patients. When integrated with conventional clinical variables, *miR-1* improved the model’s predictive accuracy. These findings suggest that *miR-1* could serve as a useful molecular marker for enhanced cardiovascular risk stratification in asymptomatic individuals with familial hypercholesterolemia.

## INTRODUCTION

Familial hypercholesterolemia (FH) is a frequent autosomal dominant disorder, primarily caused by mutations in the LDL receptor (*LDL-R*) gene, leading to disrupted cholesterol metabolism and elevated LDL-cholesterol levels since birth [1]. This disease confers an increased risk of developing premature atherosclerotic cardiovascular disease (ASCVD) [2, 3]. Previous studies have shown that imaging techniques can effectively assess the risk of atherosclerosis and plaque calcification. In this context, coronary computed tomography angiography (CTA) has been used to evaluate ASCVD risk in patients with FH enrolled in the SAFEHEART cohort (Spanish Familial Hypercholesterolemia Cohort Study) [4]. Additionally, in this same cohort, several variables associated with ASCVD risk have been identified, including age, sex, history of ASCVD, blood pressure, body mass index, smoking, and plasma levels of LDL-C and Lp(a). These factors were integrated into a risk score with potential clinical utility for assessing ASCVD risk in patients with [5]. While this approach demonstrates high sensitivity and specificity, the strong genetic basis of FH highlights the need to incorporate genetic and epigenetic markers to further enhance predictive accuracy. Previous studies have shown that adding novel biomarkers, such as proteins or lipids, to traditional risk factors (age, sex, smoking, blood pressure, etc.) significantly improves ASCVD prediction models.

MicroRNAs (miRNAs) are short non-coding RNAs (∼22 nucleotides) that regulate gene expression by binding to gene promoter regions, typically inhibiting transcription. Due to their ability to target multiple genes, and be targeted by multiple miRNAs, they form a complex regulatory network. MiRNAs have been implicated in various diseases, including cardiovascular disease, through their role in controlling disease-related biological processes[6]. In fact, recent studies have demonstrated their potential as biomarkers for the prediction of disease development and even their use as a therapeutic tool [7–10]

Based on this rationale, we hypothesize that incorporating microRNAs into clinical models could enhance the prediction of ASCVD risk. Accordingly, our objective is to identify miRNAs that, when combined with clinical variables, improve the predictive accuracy for ASCVD development in asymptomatic familial hypercholesterolemia patients from the SAFEHEART cohort.

## METHODS

### Study subjects

SAFEHEART is a multicenter, nationwide, long-term, prospective cohort study in a molecularly defined heterozygous population of patients with Familial Hypercholesterolemia (FH) in Spain. The study started in 2004 and includes index cases with a genetic diagnosis of familial hypercholesterolemia and their relatives > 15 years (with or without familial hypercholesterolemia). All cases have been molecularly characterized for FH by Next-generation sequencing that identifies functional mutations in the LDL receptor (null allele, defective allele), in the apo B receptor and in the PSCK9 gene [11]. The design and the follow-up of the SAFEHEART study have been previously described [12, 13]. For this study, data analysed were obtained between 14 January 2004 and October 2015. Subject’s ≥18 years old with genetic diagnosis of FH, without clinical coronary artery disease, on chronic and stable lipid-lowering therapy, and with a coronary CT angiography and CAC measurement using Agatston score were included [14]. For this study, homozygous FH patients were excluded. For the sample size calculation, it was taken into account that the main variable of the study was the differences in Agatson Score classification, minimum detectable proportion of patients between groups: 5%, risk alpha = 0.05, power (1-ß) =0.80, expected losses, impossibility of coronary angiography measurement or performance of Agatston Score =0.1. Based on these assumptions, a total of at least 133 participants per Agatston category x 3 possible categories = 399 patients minimum. This study was approved by the ethic committee of the Fundación Jiménez Díaz and all eligible subjects gave written informed consent. The present study base on a total population of 400 subjects diagnosed with FH, we included those patients who have clinical (especially coronary CT and Agatston score) and miRNAs determinations (n=320). The cohort SAFEHEART is registered on ClinicalTrials.gov, number NCT02693548.

### Clinical Measurements

Demographic and clinical characteristics were recorded as described elsewhere [12]. Venous blood samples were taken after a 12-hour fast. Plasma lipid profile and lipoprotein a (Lp(a)) levels were determined as previously described [15]. Because many patients were on lipid-lowering therapy (LLT) at inclusion, pretreatment LDL-C levels were estimated according to previous recommendations [16]. Hypertension was defined as systolic blood pressure >140 mm Hg and/or diastolic blood pressure >90 mm Hg on 2 measurements on 2 different days or need of antihypertensive drugs. DNA was isolated from whole blood, and the genetic diagnosis of FH was carried out as previously described [17]. Mutations were classified as receptor negative or receptor defective, depending on their functional class. Mutations without a functional class in the literature were classified as unclassified mutations [18]. The potency of LLT was calculated as reported elsewhere with a modification to include the effect of ezetimibe [19, 20]. Ezetimibe was considered to decrease LDL-C by 15%, and this effect was added to the statin effect when appropriate [19]. Serum LDL-C levels were calculated using the Friedewald formula and LDL burden was calculated as the median of the observed LDL-cholesterol level multiplied by the number of years, and adjusted for gender [21].

### Coronary artery calcification

Coronary artery calcification (CAC) was assessed by coronary CT angiography as previously described for the SAFEHEART cohort [22]. For the study of ASCVD prediction using miRNAs, the population included in the present study was categorized into two groups based on the Agatston score. Subjects with low and medium risk (Agatston score < 299) and high risk (Agatston score > 300) were grouped together.

### Isolation of circulating miRNAs from plasma samples

Venous blood from the 320 subjects molecularly diagnosed with familial hypercholesterolaemia was collected on a single visit in tubes containing EDTA, and centrifuged at 2.000 x g for 10 min to separate the plasma from the blood cells. RNA isolation was carried out from plasma samples, as previously described by Jimenez-Lucena et al [8].

### cDNA synthesis and circulating miRNAs levels by real-time PCR

The cDNA synthesis was carried out using the TaqMan MicroRNA Reverse Transcription Kit (Life Technologies – Thermofisher Scientific, Carlsbad, CA, USA) following the manufacturer’s instructions, as previously described in our group by Jimenez-Lucena et al [8]. The circulating miRNAs study was carried out on 28 miRNAs in duplicate. The 28 miRNAs were selected based on a bibliographic search according to their association with lipid metabolism, endothelial dysfunction and cardiovascular disease (**Supplementary Table 1**). We measured the levels of miRNAs using the OpenArray® platform (Life Technologies – Thermofisher Scientific, Carlsbad, CA, USA) following the manufacturer’s instructions. The relative expression data was analyzed using OpenArray® Real-Time qPCR Analysis Software (Life Technologies – Thermofisher Scientific, Carlsbad, CA, USA) and the normalization method has been previously described [8].

### Statistical analyses

To analyse the differences in biochemical characteristics and miRNAs levels between subjects carrying the null allele and the defective allele for the *LDL-R* gene, a ONE-WAY ANOVA analysis was performed. In the same way, the analysis of the baseline characteristics of the subjects included in the study was carried out according to the Agatston score categories (<299 and >300). The same categorization was used for the assessment of ASCVD risk using ROC models. The analyses described above were carried out using the statistical software SPSS 21. p < 0.05 was considered statistically significant. ASCVD risk for an individual with FH was estimated using the SAFEHEART-RE modified risk equation (gender, age, diagnosis of arterial hypertension, smoking activity, body mass index, LDL-burden, Lp(a)) [23]. Given that ASCVD is the study variable and that none of the subjects included in the present study had a family history of ASCVD, this variable was excluded from the calculation of the score. ROC curve analyses were carried out using R software. ROC curves were constructed using the pROC package and using the `glm` function, evaluating sensitivity, specificity, accuracy, threshold and area under the curve of each model. De Long’s test was performed using the `roc.test` function evaluating the z-value and p-value between models. p < 0.05 was considered statistically significant.

### Identification of cellular pathways and genes regulated by miRNAs of interest

In order to understand the role of miRNAs in the regulation of biological processes related to familial hypercholesterolemia and ASCVD, we conducted an analysis with the bioinformatics tool MicroRNA Enrichment Turned Network (MIENTURNET) based on miRTarBase database [24]. The miRNA identified with potential of biomarker in our study were included in the analysis. The filter results setup was using a threshold for the minimum number of miRNA-target interactions = 1 and the set a threshold for the adjusted p-value (FDR) = 0.1. We removed pathways which are specific to cancer and infectious diseases and obtained the biological pathways regulated by miRNAs of interest.

## RESULTS

### Characteristics of the subjects included in the present study

The general characteristics of the population included in this study are described in **Table 1**. Participants were classified based on whether they carried a defective or null allele of the *LDL-R* gene. Based on this classification, most participants were carriers of the defective allele (n = 194), compared to those with the null allele (n = 126) (p < 0.001; **Figure 1A**). However, no significant differences were observed in the baseline characteristics of the subjects included in the study (**Table 2**). Additionally, participants were categorized based on their Agatston calcification score. Significant differences were found in age and BMI, with individuals with score > 300 exhibiting higher values compared to those with score < 299 (p<0.001 and p=0.0175, respectively) (**Table 3**).

**Figure 1.**
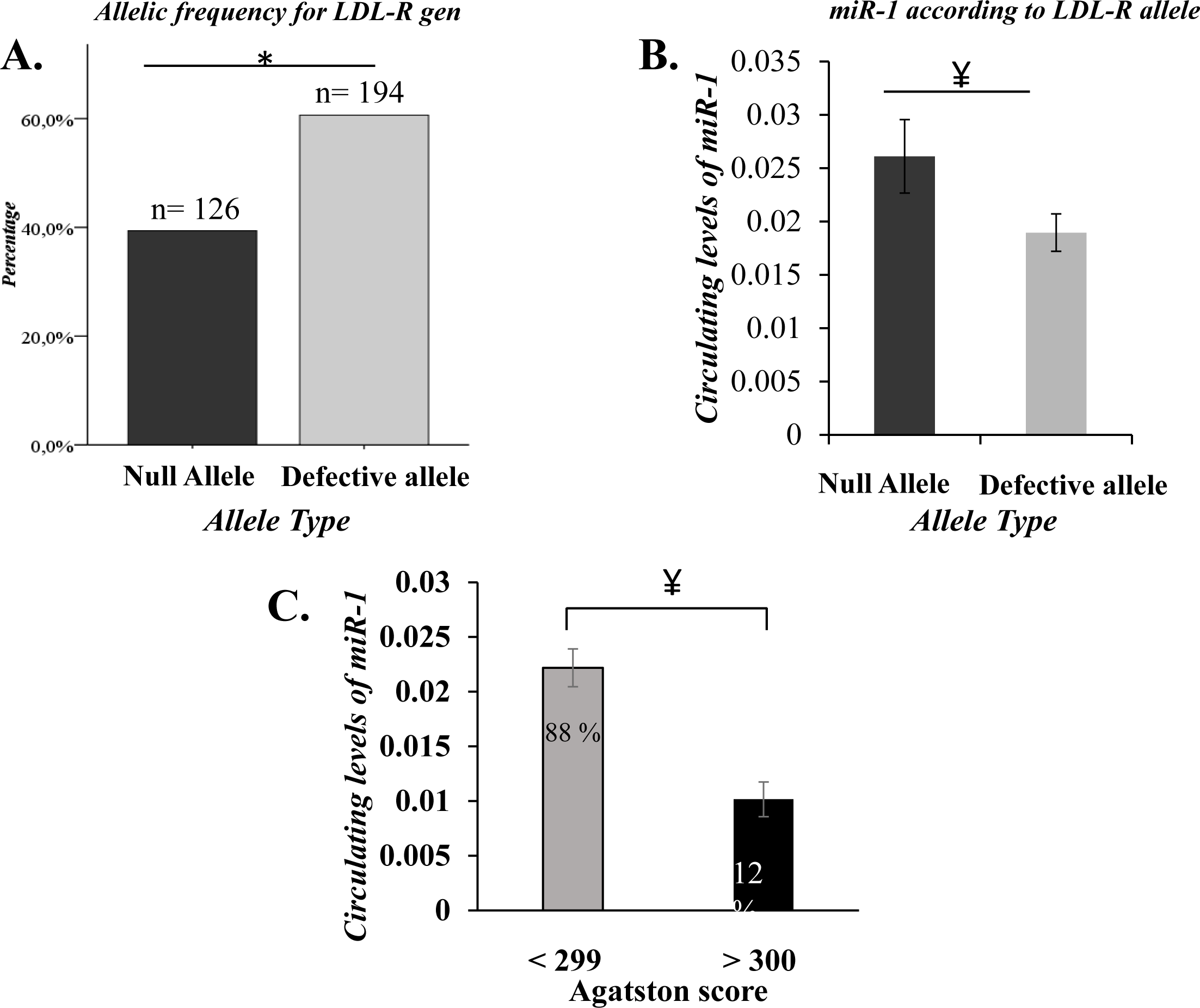
Genetics and miRNAs relationship in the SAFEHEART Registry. **A.** Allelic frequency in the *LDL-R* gen, data represent the percentage of subjects according to allele frequency for *LDL-R* gene. **B.** Circulating *miR-1* levels according to allelic frequency of *LDL-R* gen, * Chi square test p < 0.05. ¥ ONE-WAY Anova test p < 0.05. **C.** Circulating levels of *miR-1* according to the Agatston score. Data represent the mean value ± standard error. The analysis corresponds to a one-way ANOVA. * p< 0.05. Inside each bar, the percentage of the population that falls into the corresponding Agatston score category is shown. Analyses were performed using SPSS 21.

**Table 1.**
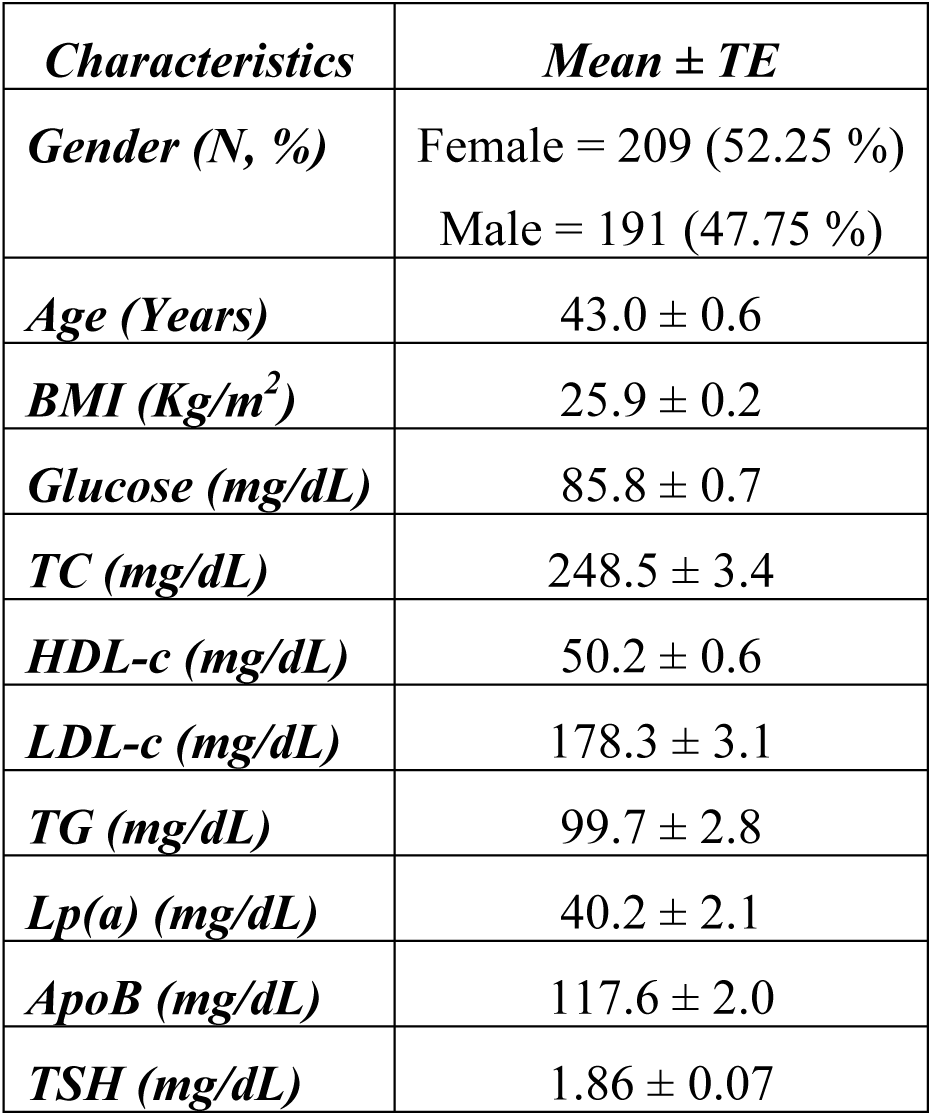
Descriptive of the population included in the present study. Data represent the mean ± typical error (TE). TC: Total cholesterol; HDL-c: High-density cholesterol; LDL-c: Low-density cholesterol; TG: Triglycerides; Lp(a): Lipoprotein A; ApoAI: Apolipoprotein AI; ApoB: Apolipoprotein B; TSH: Thyroid stimulating hormone.

| *Characteristics* | *Mean ± TE* |
| --- | --- |
| *Gender (N, %)* | Female = 209 (52.25 %)<br>Male = 191 (47.75 %) |
| *Age (Years)* | 43.0 ± 0.6 |
| *BMI (Kg/m<sup>2</sup>)* | 25.9 ± 0.2 |
| *Glucose (mg/dL)* | 85.8 ± 0.7 |
| *TC (mg/dL)* | 248.5 ± 3.4 |
| *HDL-c (mg/dL)* | 50.2 ± 0.6 |
| *LDL-c (mg/dL)* | 178.3 ± 3.1 |
| *TG (mg/dL)* | 99.7 ± 2.8 |
| *Lp(a) (mg/dL)* | 40.2 ± 2.1 |
| *ApoB (mg/dL)* | 117.6 ± 2.0 |
| *TSH (mg/dL)* | 1.86 ± 0.07 |

**Table 2.** Characteristics of the subjects included in the present study according the allelic frequency for the *LDL-R* gen. Data represent the mean ± typical error (TE). The data correspond to ONE WAY Anova analysis with SPSS 21. p < 0.05 was considered as statistically significant. TC: Total cholesterol; HDL-c: High-density cholesterol; LDL-c: Low-density cholesterol; TG: Triglycerides; Lp(a): Lipoprotein A; ApoB: Apolipoprotein B; TSH: Thyroid stimulating hormone.

|  | *Null Allele* | *Defective Allele* | *p-value* |
| --- | --- | --- | --- |
| *N* | 126 | 194 |  |
| *TC (mg/dL)* | 248.5 ± 6.2 | 255.2 ± 5.1 | 0.4069 |
| *TG (mg/dL)* | 96.4 ± 4.0 | 104.7 ± 4.7 | 0.2152 |
| *HDL-c (mg/dL)* | 49.4 ± 1.1 | 50.0 ± 1.0 | 0.6954 |
| *LDL-c (mg/dL)* | 179.8 ± 5.6 | 184.3 ± 4.7 | 0.5458 |
| *ApoB (mg/dL)* | 120.6 ± 3.4 | 119.3 ± 2.9 | 0.7667 |
| *Lp(a) (mg/dL)* | 44.5 ± 4.1 | 37.2 ± 2.7 | 0.1233 |
| *TSH (mg/dL)* | 1.7 ± 0.1 | 1.9 ± 0.1 | 0.2599 |
| *Glucosa (mg/dL)* | 85.8 ± 1.2 | 86.0 ± 1.0 | 0.9191 |
| *Age (Years)* | 42.0 ± 1.1 | 42.9 ± 0.8 | 0.4892 |
| *BMI (Kg/m2)* | 25.7 ± 0.4 | 26.0 ± 0.3 | 0.5718 |

**Table 3.** Characteristics of the subjects included in the present study according to the Agatston score. Data represent the mean ± typical error (TE). The data correspond to ONE WAY Anova analysis with SPSS 21. * p < 0.05 was considered as statistically significant. TC: Total cholesterol; HDL-c: High-density cholesterol; LDL-c: Low-density cholesterol; TG: Triglycerides; Lp(a): Lipoprotein A; ApoB: Apolipoprotein B; TSH: Thyroid stimulating hormone.

|  | *Agatston < 299* | *Agatston > 300* | *p-value* |
| --- | --- | --- | --- |
| *TC (mg/dL)* | 249.48 ± 3.89 | 256.94 ± 11.14 | 0.5081 |
| *TG (mg/dL)* | 100.60 ± 3.38 | 99.08 ± 5.30 | 0.8697 |
| *HDL-c (mg/dL)* | 50.05 ± 0.72 | 49.87 ± 1.94 | 0.9287 |
| *LDL-c (mg/dL)* | 179.27 ± 3.57 | 187.13 ± 10.04 | 0.4463 |
| *ApoB (mg/dL)* | 117.18 ± 2.27 | 130.00 ± 5.45 | 0.0541 |
| *Lp(a) (mg/dL)* | 39.40 ± 2.32 | 53.46 ± 8.22 | 0.0503 |
| *TSH (mg/dL)* | 1.82 ± 0.07 | 2.13 ± 0.43 | 0.1881 |
| *Glucosa (mg/dL)* | 85.82 ± 0.78 | 89.06 ± 2.06 | 0.1738 |
| *Age (Years)* | 41.75 ± 0.63 | 50.34 ± 1.19 | < 0.001* |
| *BMI (Kg/m<sup>2</sup>)* | 25.66 ± 0.25 | 27.30 ± 0.49 | 0.0175* |

### Circulating miRNAs levels according to the Agatston score

Following the assessment of circulating miRNA levels, 15 miRNAs failed to amplify in at least 80% of the samples and were excluded from further analysis. *miR-423-5p*, identified as the most stable across all samples, was selected for data normalization. Thus, our study had a database with 12 miRNAs. We analysed the circulating miRNA levels in relation to the Agatston score and found that only *miR-1* showed significant differences between groups (**Supplementary Table 2**), while the remaining miRNAs did not. Additionally, subjects carrying the null LDL-R allele exhibited higher circulating levels of *miR-1* compared to those with the defective allele (p=0.042)(**Figure 1B**). Circulating *miR-1* levels were also analysed according to the Agatston score. Individuals at lower cardiovascular risk (score < 299) exhibited significantly higher *miR-1* levels compared to those at higher risk (score >300) (p=0.011)(**Figure 1C**). In addition, no statistically significant differences were observed in circulating *miR-1* levels between male and female groups.

### Analyses of receiver operating curves (ROC) based on variables included in the SAFEHEART-RE and miRNAs

To evaluate the potential of miRNAs in assessing ASCVD risk among individuals with familial hypercholesterolemia, we performed ROC curve analyses using the Agatston calcification score as the grouping variable (<299 vs. >300). The initial model included the variables from the SAFEHEART-RE score—gender, age, hypertension diagnosis, smoking status, body mass index, LDL burden, and Lp(a)— yielding an AUC of 0.846 (95% CI: 0.789–0.902) (sensitivity = 0.77; specificity = 0.77) (**Figure 2A**). Subsequently, a second model was constructed by incorporating *miR-1* levels into the variables included in the initial model. The second model showed an AUC = 0.878 (CI: 0.809 - 0.937) (sensitivity = 0.94; specificity = 0.66) (**Figure 2B**). In addition, we conducted a De Long test to compare the two models and observed a significant difference between them, a z-value = -2.7886 and p-value = 0.0052 (**Figure 2C**).

**Figure 2.**
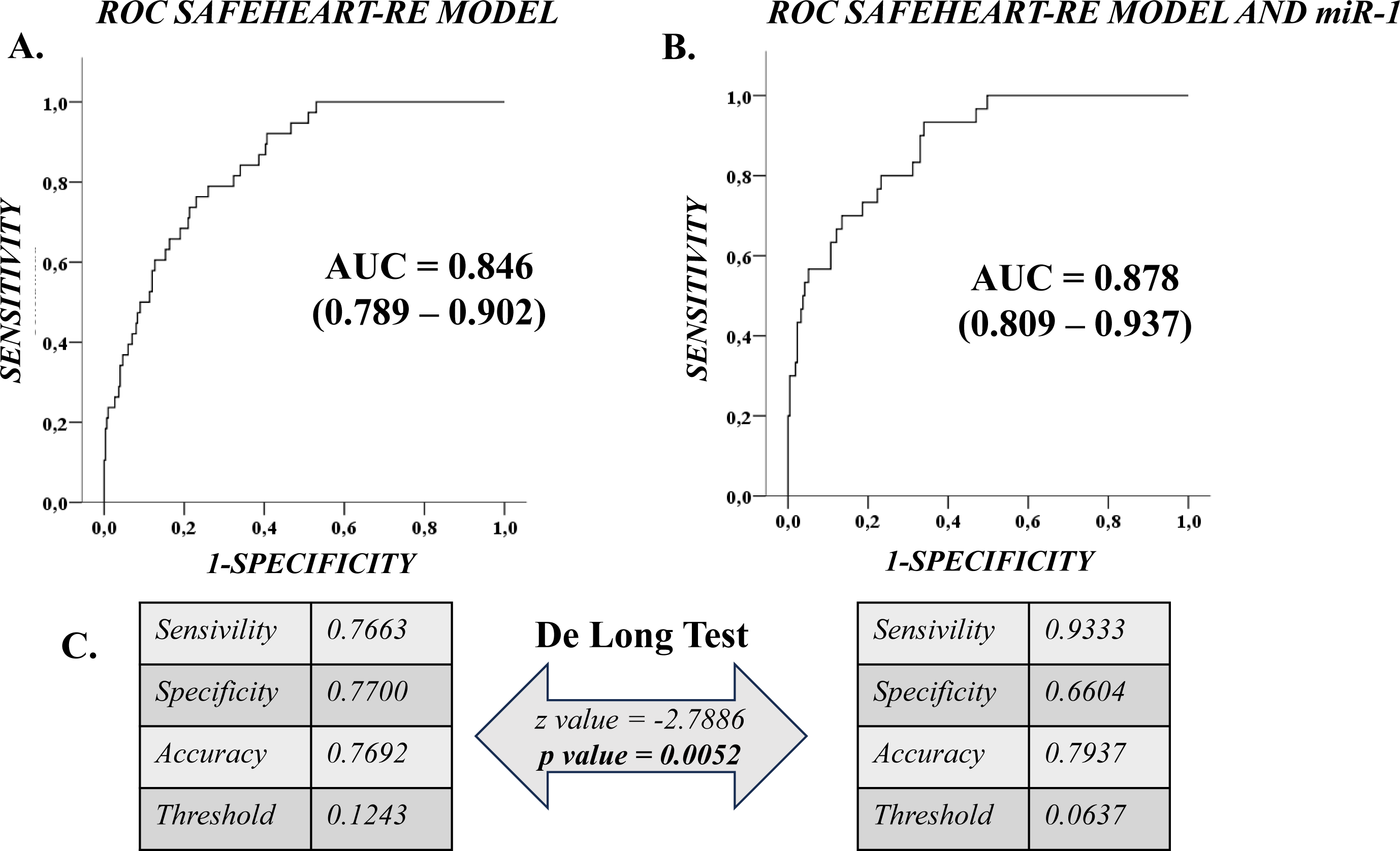
ROC curve analyses. **A.** ROC model based on SAFEHEART-RE variables (gender, age, high blood pressure, history of ASCVD, active smoking, body mass index, LDL-burden, Lp(a). **B.** ROC model SAFEHEART-RE variables and added to *miR-1* levels. Analyses were carried out using R software. ROC curves were constructed using the pROC package and using the `glm` function, evaluating sensitivity, specificity, accuracy, threshold and area under the curve of each model. **C.** De Long’s test was performed using the `roc.test` function evaluating the z-value and p-value between the dos models. p < 0.05 was considered statistically significant. AUC = Area under curve.

### Enrichment analysis of genes and pathways regulated by *miR-1*

We carried out an analysis using the MIELTURNET platform to identify the biological processes and genes regulated by the miRNA of interest. Thus, based on the lowest FDR value and showing at least 1 interaction with *miR-1*, the top 10 genes regulated by this miRNA were *ADAMTSL4, AIM2, BCKDHB, C6orf118, CAVIN2, CCDC102A, CEBPZ, CETN3, CLEC1A* and *DEFB125* (FDR = 0.0064; p= 0.0004; in all genes). Additionally, reactome pathway analysis identified five pathways involving *miR-1*, with the most relevant to atherosclerotic plaque formation being platelet activation, signalling and aggregation; nuclear receptor signalling; and FGFR4-related disease signalling (**Figure 3**).

**Figure 3.**
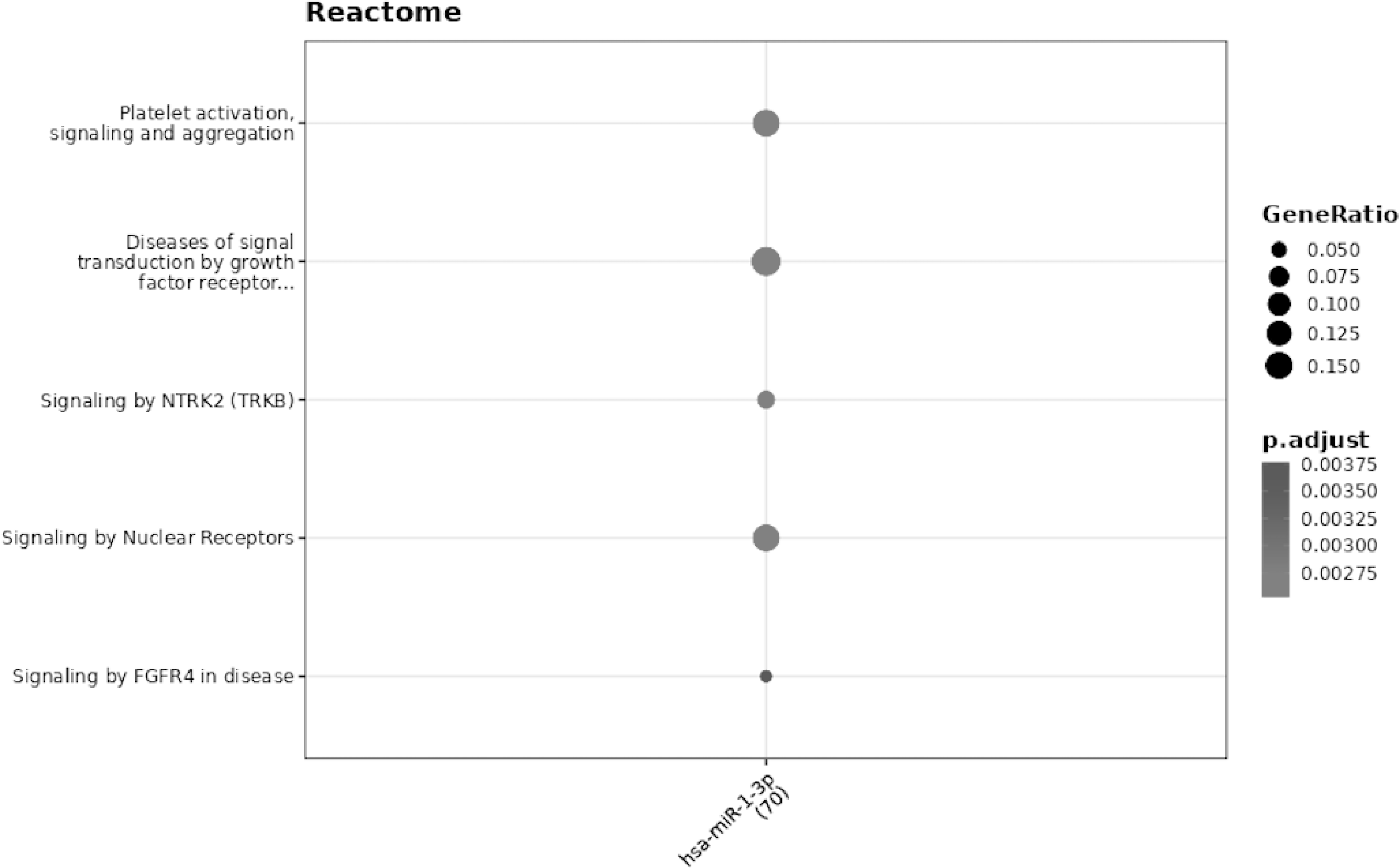
Enrichment analysis of pathways regulated by *miR-1*. The analysis was performed by MIENTURNET based on miRTarBase database.

## DISCUSSION

Our study, based on the SAFEHEART cohort (Spanish Familial Hypercholesterolemia Cohort Study), demonstrates that adding *miR-1* to traditional clinical variables significantly improves ASCVD risk assessment by 3.8% in asymptomatic ASCVD FH patients, as confirmed by the DeLong test. Previous studies analyzing patients from the same cohort, showed elevated plasma levels of *miR-6821-5p*, which were associated with increased expression of the calcification-related genes *SPP1* and *BMP2*, as well as the presence of subclinical calcified atherosclerotic plaques [25]. In another study, increased circulating expression of *miR-604, miR-652-5p* and *miR-4451*, as well as reduced expression of *miR-3140-3p*, *miR-550a-5p* and *miR-363-3p* was observed in subjects with Familial Hypercholesterolemia (FH) compared to healthy subjects [26]. In turn, Costa da Freitas *et al*, demonstrated in 33 subjects with FH patients, increased levels of *miR-122-5p* compared to a group of 41 controls[27]. While *miR-1* has not been previously reported as differentially expressed in individuals with FH, it has been associated with arterial markers of atherosclerosis through its involvement in crucial pathways such as endothelial function and angiogenesis [28], apoptosis [29, 30] and differentiation of cardiac myocytes [29, 31].

The primary manifestation of FH is elevated LDL cholesterol levels since birth, which significantly increase the risk of ASCVD. This risk can be assessed using imaging tools such as coronary artery calcification scoring by angiography (Agatston score), or through clinical risk scores like the SAFEHEART-RE score [5]. In this regards, recent studies have shown that a high coronary calcium CT score is associated with older age and is more prevalent in men than in women, regardless of age, even among healthy individuals [32]. However, considering the genetic basis of familial hypercholesterolaemia, it is essential to incorporate genetic or epigenetic factors to enhance the assessment of ASCVD.

Interestingly, our findings revealed elevated levels of *miR-1* in carriers of null *LDL-R* alleles compared to those with defective variants. Similarly, *miR-1* levels were found to be higher in subjects with an Agatston score below 299 compared to those with a score above 300. Furthermore, ROC curve analysis demonstrated that ASCVD risk prediction based on the SAFEHEART-RE score—which includes clinical variables such as sex, age, arterial hypertension, smoking status, body mass index, LDL burden, and Lp(a) levels—achieved an AUC of 0.846. Notably, when *miR-1* was incorporated into the model, the predictive performance improved, yielding an AUC of 0.878. As previously mentioned, the DeLong test confirmed that the difference between the two models was statistically significant (z=-2.79; p=0.005). Finally, gene and pathway analysis revealed that *miR-1* regulates several target genes, including *AIM2, CAVIN2, and CEBPZ*, as well as biological processes such as: platelet activation, cellular aggregation, nuclear receptor signalling, and FGFR4 signalling pathways implicated in disease.

In this sense, our *in silico* analysis, revealed that *miR-1* interacts with (and potentially inhibits) at least 10 target genes. Notably, one of these genes, *AIM2*, has been previously identified as a key sensor of cholesterol-dependent inflammasome activation, implicating it in the initiation of proinflammatory responses triggered by elevated cholesterol level [33, 34]. In turn, caveolin 2 is encoded by the *CAVIN2* gene, and is a major protein component of the cholesterol- and glycosphingolipid-rich flask-shaped invaginations of plasma membrane caveolae [35]. Previous studies have shown that caveolin 2 is rapidly degraded by cholesterol depletion leading to caveolae collapse [36]. This indicates that inhibition of *CAVIN2* by *miR-1* may be associated with dysregulation of cholesterol metabolism. Moreover, the *CEBPZ* gene belongs to the CCAAT/enhancer binding proteins (C/EBPs) family, which are transcriptional factors involved in the activation of many inflammatory mediators. Additionally, a previous study in a KO mouse model for the LDL receptor (*LDLR*-/-), demonstrated that silencing of C/EBPε in mouse macrophages may have the ability to decrease the development of atherosclerosis and change lipid metabolism [37].

In our cohort, we analysed circulating levels of *miR-1* in men and women and found no significant differences. Previous studies have shown that the burden and risk of ASCVD are significantly lower in women than in men with familial hypercholesterolaemia [38]. However, higher *miR-1* levels were observed in individuals at lower risk of atherosclerotic cardiovascular disease, as indicated by Agatston scores below 299. These findings suggest that *miR-1* may play an anti-inflammatory and cardioprotective role. This is supported by the fact that previous studies suggest that *miR-1* expression of the above-mentioned target genes would downregulate proinflammatory activity and induce changes in cholesterol metabolism, suggesting a lower risk of atherosclerosis development. However, further research is needed to elucidate the underlying biological mechanisms that support this potential cardioprotective role of *miR-1* in atherosclerosis among patients with FH.

This study has certain limitations, the most significant being the targeted analysis of 28 miRNAs selected from the literature based on their known associations with lipid metabolism, endothelial dysfunction, and cardiovascular disease. As a result, other miRNAs not previously linked to these processes—and potentially of greater relevance—may have been overlooked.

In conclusion, our study identified *miR-1* as a potential biomarker which, when combined with clinical variables, could improve the assessment of ASCVD risk in asymptomatic individuals with FH.

## Data Availability

The availability of data from the present study is subject to collaboration with the research group and authorisation from the Familial Hypercholesterolaemia Foundation. Collaborations are open to Biomedical Institutions, always after an accepted proposal for a scientific work.

## Non-standard Abbreviations and Acronyms

ASCVD: Atherosclerotic cardiovascular disease
AUC: Area under curve
CAC: Coronary Artery Calcification
CTA: Computed tomography angiography
FH: Familial hypercholesterolemia
LDL-R: LDL receptor
LLT: Lipid-lowering therapy
Lp(a): Lipoproteina (a)
miRNA: MicroRNA
ROC: Receiver operating curve
SAFEHEART: Spanish Familial Hypercholesterolemia Cohort Study

## ACKNOWLEDGMENTS

The authors thank the Spanish Familial Hypercholesterolemia Foundation for assistance in the recruitment and follow-up of participants and the families with familial hypercholesterolemia for their valuable contribution and willingness to participate.

## Highlights

1. Study finds *miR-1* improves ASCVD risk prediction in FH patients.
2. *miR-1* emerges as a potential biomarker for early ASCVD detection.
3. Use of miRNAs for ASCVD prediction in the SAFEHEART cohort

